# Serological prevalence of SARS-CoV-2 infection and associated factors in healthcare workers in a “non-COVID” hospital in Mexico City

**DOI:** 10.1101/2020.11.30.20241331

**Authors:** Esteban Cruz-Arenas, Elizabeth Cabrera-Ruiz, Sara Laguna-Barcenas, Claudia A. Colin-Castro, Tatiana Chavez, Rafael Franco-Cendejas, Clemente Ibarra, Javier Perez-Orive

**Affiliations:** Instituto Nacional de Rehabilitacion “Luis Guillermo Ibarra Ibarra”, Mexico City, Mexico; Epidemiological Vigilance Unit, Instituto Nacional de Rehabilitacion “Luis Guillermo Ibarra Ibarra”, Mexico City, Mexico; Infectious Diseases Division, Instituto Nacional de Rehabilitacion “Luis Guillermo Ibarra Ibarra”, Mexico City, Mexico

**Author notes:** Corresponding author: Javier Perez-Orive, Instituto Nacional de Rehabilitacion “Luis Guillermo Ibarra Ibarra”, Calzada Mexico-Xochimilco 289, Mexico City 14389, Mexico.

## Abstract

In spite of high mortality from COVID-19, in Mexico the number of confirmed cases and diagnostic tests per million population are lower than for other comparable countries, which leads to uncertainty about the actual extent of the pandemic. In Mexico City, healthcare workers represent an important fraction of individuals with SARS-CoV-2 infection. This work aims to estimate the frequency of antibodies to SARS-CoV-2 and identify associated factors in healthcare workers at a large hospital in Mexico City. We conducted a serological survey in a non-COVID national referral teaching hospital. We selected a representative sample of 300 individuals. Blood samples were collected and questionnaires were applied between August 10^th^ and September 9^th^, 2020. ELISA results indicate a serological prevalence of SARS-CoV-2 infection of 13.0%. Working in the janitorial and security groups, having an educational level below a university degree, and living with a larger number of people, were also identified as sociodemographic factors that increase the risk of having SARS-CoV-2 infection. Thus, less favored socioeconomic groups are at significantly higher risk of experiencing SARS-CoV-2 infection. Even in healthcare workers there is still a majority of individuals that are seronegative, and thus the risk of continued epidemic waves and mortality remains high.

## INTRODUCTION

Severe acute respiratory syndrome coronavirus 2 (SARS-CoV-2) was identified in Wuhan, China at the end of 2019 and is the cause of Coronavirus disease 2019 (COVID-19). SARS-CoV-2 spread globally in early 2020, with the first cases reported outside China on January 13^th^ and 29^th^ in South-East Asia and the Eastern Mediterranean and on February 21^st^, 22^nd^ and 25^th^ in the Americas, Europe and Africa, respectively^1^. The dynamics of infections have been different in each region. Since May the Americas have occupied the first place worldwide in the number of COVID-19 deaths reported (as of November 6^th^, 650,705 deaths from a global death toll of 1,231,017)^1^. Countries in the Americas occupy three of the first four places in number of deaths: the United States of America (USA) with 232,166, Brazil with 161,106 and Mexico with 93,772 ^1^. Quantified as number of deaths per million population, Brazil occupies the 6^th^ place globally with 757.9, Mexico the 9^th^ with 723.1, and the USA the 11th with 701.4. However, the number of reported cases per million inhabitants in Mexico (7,318.8) is much lower than that of the USA and Brazil (with 28,362.2 and 26,298.7, respectively)^1^. The number and criteria for conducting real time polymerase chain reaction (RT-PCR) tests in each country have also been different. As of October 30^th^, the estimates are 16.1 tests per million inhabitants for Mexico, compared to 456.0 for the United States and 30.2 for Brazil^2^. This smaller number of RT-PCR tests in Mexico is related to a very high positivity rate of tests that was 41.3% on September 9^th 3^. This leads to much uncertainty about the actual number of COVID-19 infections that have occurred in countries with low numbers of tests per million population.

In Mexico the first case was reported on February 28^th^ ^1^. On March 31^st^ extraordinary government measures to attend the health emergency were taken: social distancing; closing of all “non-essential” economic activity; the conversion of hospitals into exclusive units for the management of pandemic patients, i.e. “COVID hospitals”, and “non-COVID hospitals” which would focus on non-COVID-19 patients requiring other medical care; designation of the sentinel model of epidemiological surveillance^1^ and focusing RT-PCR tests on people with symptoms^3^. There was a large impact to the economy with Gross Domestic Product decrease in the second quarter of 18.7%, compared to the same quarter of the previous year^4^. Healthcare workers (HCW) represent an important fraction of individuals with SARS-CoV-2 infection in Mexico City^5^. Our institution is a public national referral teaching hospital with 235 beds located in Mexico City and was designated by the federal health authorities as a non-COVID hospital. Hospitals designated as non-COVID delayed elective surgeries focusing on urgencies. Our institution became one of the few public hospitals in Mexico City attending trauma patients. Upon arrival at the emergency room, all patients and relatives were asked about possible COVID-19 contacts, respiratory symptoms and comorbidities in order to identify infection risk factors. During their stay at the emergency room patients were clinically evaluated and then discharged to the next filter where they were swabbed for the RT-PCR test. Non-COVID hospitals are not prepared to treat COVID-19 patients, thus if any patient was identified as infected he or she was referred to a COVID hospital if their symptoms required hospital attention, otherwise they were directed to self-isolate at home and asked to come back once the infection subsided.^3^

Accurate epidemiological information is required to support public health actions aimed at limiting contagion and loss of life, while minimizing negative economic impact to the population^1^. With this purpose, serological surveys estimating SARS-CoV-2 infection have been conducted in many places globally, finding for instance seroprevalence levels of 4.1 %, in April in Los Angeles, California^6^, 6.9% in March^7^ and 22.7% in April^8^ in New York City, and 11.5% in May in Madrid, Spain^9^. Other studies of seroprevalence quantified in the same city on several occasions find in Geneva, Switzerland for 5 consecutive weeks 4.8%, 8.5%, 10.9%, 6.6% and 10.8%, respectively^10^, and in London, England for the 13^th^, 15^th^, and 18^th^weeks of 2020 1.5%, 12.3% and 17.5%, respectively^11^. Studies carried out in HCWs in Denmark find seroprevalence of 29.7% for emergency departments, and 2.2% for departments with limited or no patient contact^12^; in Sweden 19.1% for a large acute care hospital, of which 21.0% for HCW with direct patient contact, and 8.5% for those without patient contact^13^; in London, England 34.7% in a clinical environment with prolonged direct contact with patients, and 22.6% in a non-clinical environment with minimal or no patient contact^14^; in Spain 10.2% for HCW in general^9^; in the New York City area 13.1% for nurses, 8.7% for physicians, 12.6% for administrative and clerical, and 20.9% for service-maintenance personnel^15^; in a tertiary care Center in Belgium 6.4%^16^. In Mexico there are yet no published seroprevalence studies that allow for a more accurate description of the prevalence of SARS-CoV-2 infections in a Mexican population.^17^Regarding the characteristics of antibody responses, a cohort study^18^ showed SARS-CoV-2 specific neutralizing antibodies in 94% of 175 patients with clinically mild COVID-19 within 2 weeks after the onset of symptoms (30% of patients with very low level of neutralizing antibodies, 17% medium-low, 39% medium-high, and 14% high). Other studies have shown positive immunoglobulin G (IgG) antibody tests of 100% for 285 patients with COVID-19 within 19 days after symptom onset^19^; and 94.3%, and 79.8%, for immunoglobulin M (IgM) and IgG, respectively, by day 15 after onset^20^. A study following 19 patients with positive SARS-CoV-2 antibody tests found that 60 days later 11 (58%) of these patients became seronegative, while only 8 (42%) remained above the seropositivity threshold.^21^

This work provides serological data from a non-COVID national referral teaching hospital. Its main objective is to estimate the frequency of antibodies to SARS-CoV-2 in HCW at a large non-COVID hospital in Mexico City. By conducting questionnaires with the study participants, it also allows us to identify sociodemographic and other factors associated with SARS-CoV-2 infections, as well as potential protection or risk factors for this population. Given the limited availability of COVID-19 prevalence data in Mexico^17^, the information presented here can be useful for the planning of public health measures, including for institutions with similarities to this one.

## RESULTS

### Study population

The sample included a total of 300 participants, all of whom responded to questionnaires and had Lateral Flow Assay (LFA) tests applied. Enzyme-Linked Immunosorbent Assay (ELISA) tests were conducted on 299 participants, since in one subject it was not possible to extract enough blood volume for the ELISA test, but only for the LFA test. As observed in Table 1, the largest work group strata were administrative (21.4%), medical personnel (19.4%), and nursing (18.7%). The majority of participants work in the morning shift (60.2%), are female (65.2%; in line with the overall population working in the hospital), single (49.8%), belong to the 30-44 age group (41.5%), and reported having a university bachelor’s degree or higher (72.9%), as well as high levels of patient contact (69.6%).

**Table 1.**
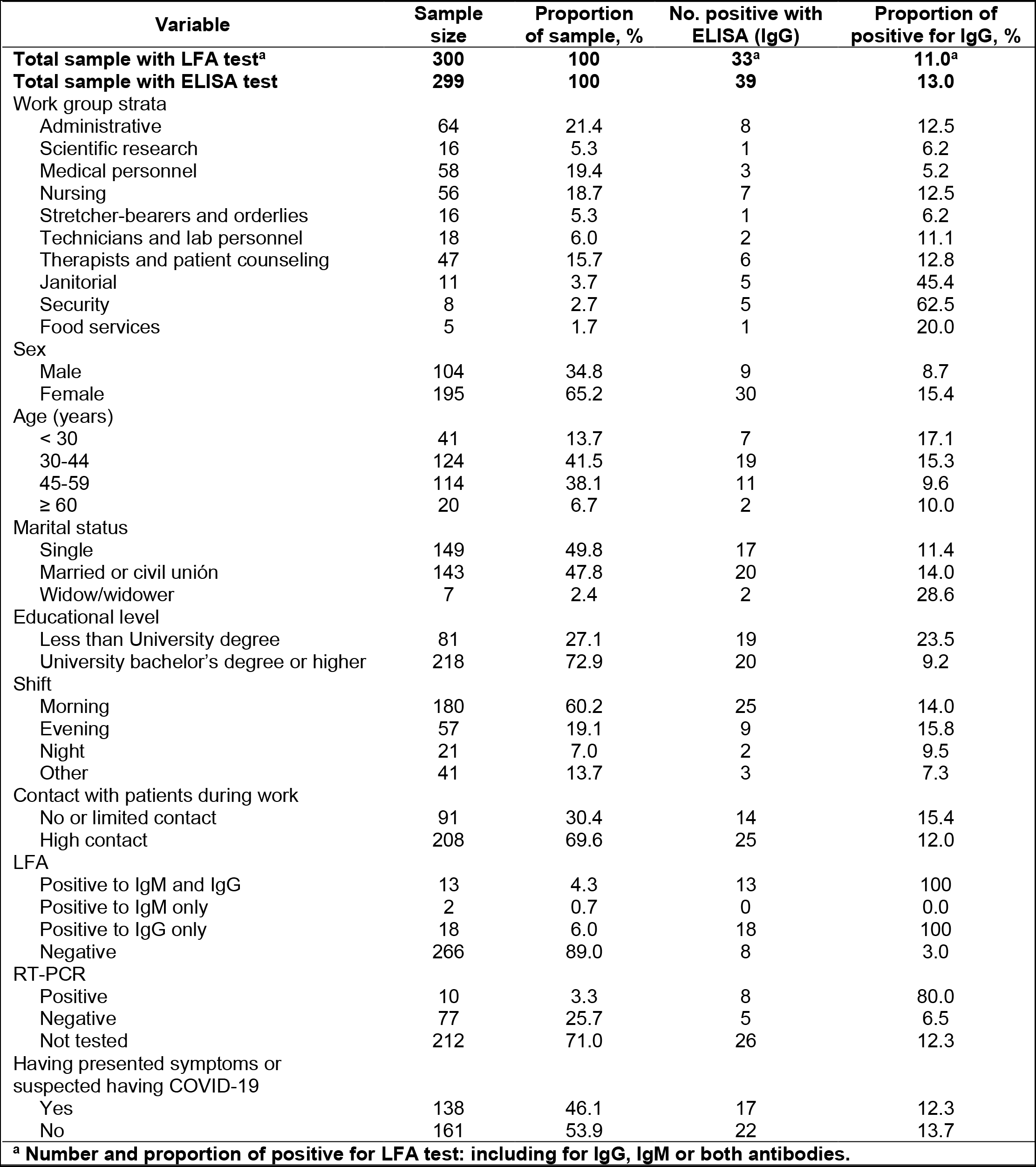
General characteristics of the study population in relationship to the categorical ELISA test results.

| Variable | Sample size | Proportion of sample, % | No. positive with ELISA (IgG) | Proportion of positive for IgG, % |
| --- | --- | --- | --- | --- |
| **Total sample with LFA test<sup>a</sup>** | **300** | **100** | **33<sup>a</sup>** | **11.0<sup>a</sup>** |
| **Total sample with ELISA test** | **299** | **100** | **39** | **13.0** |
| Work group strata |  |  |  |  |
| Administrative | 64 | 21.4 | 8 | 12.5 |
| Scientific research | 16 | 5.3 | 1 | 6.2 |
| Medical personnel | 58 | 19.4 | 3 | 5.2 |
| Nursing | 56 | 18.7 | 7 | 12.5 |
| Stretcher-bearers and orderlies | 16 | 5.3 | 1 | 6.2 |
| Technicians and lab personnel | 18 | 6.0 | 2 | 11.1 |
| Therapists and patient counseling | 47 | 15.7 | 6 | 12.8 |
| Janitorial | 11 | 3.7 | 5 | 45.4 |
| Security | 8 | 2.7 | 5 | 62.5 |
| Food services | 5 | 1.7 | 1 | 20.0 |
| Sex |  |  |  |  |
| Male | 104 | 34.8 | 9 | 8.7 |
| Female | 195 | 65.2 | 30 | 15.4 |
| Age (years) |  |  |  |  |
| < 30 | 41 | 13.7 | 7 | 17.1 |
| 30-44 | 124 | 41.5 | 19 | 15.3 |
| 45-59 | 114 | 38.1 | 11 | 9.6 |
| ≥ 60 | 20 | 6.7 | 2 | 10.0 |
| Marital status |  |  |  |  |
| Single | 149 | 49.8 | 17 | 11.4 |
| Married or civil unión | 143 | 47.8 | 20 | 14.0 |
| Widow/widower | 7 | 2.4 | 2 | 28.6 |
| Educational level |  |  |  |  |
| Less than University degree | 81 | 27.1 | 19 | 23.5 |
| University bachelor's degree or higher | 218 | 72.9 | 20 | 9.2 |
| Shift |  |  |  |  |
| Morning | 180 | 60.2 | 25 | 14.0 |
| Evening | 57 | 19.1 | 9 | 15.8 |
| Night | 21 | 7.0 | 2 | 9.5 |
| Other | 41 | 13.7 | 3 | 7.3 |
| Contact with patients during work |  |  |  |  |
| No or limited contact | 91 | 30.4 | 14 | 15.4 |
| High contact | 208 | 69.6 | 25 | 12.0 |
| LFA |  |  |  |  |
| Positive to IgM and IgG | 13 | 4.3 | 13 | 100 |
| Positive to IgM only | 2 | 0.7 | 0 | 0.0 |
| Positive to IgG only | 18 | 6.0 | 18 | 100 |
| Negative | 266 | 89.0 | 8 | 3.0 |
| RT-PCR |  |  |  |  |
| Positive | 10 | 3.3 | 8 | 80.0 |
| Negative | 77 | 25.7 | 5 | 6.5 |
| Not tested | 212 | 71.0 | 26 | 12.3 |
| Having presented symptoms or suspected having COVID-19 |  |  |  |  |
| Yes | 138 | 46.1 | 17 | 12.3 |
| No | 161 | 53.9 | 22 | 13.7 |

<sup>a</sup> Number and proportion of positive for LFA test: including for IgG, IgM or both antibodies.

### Prevalence of SARS-CoV-2 antibodies

The LFA tests resulted in a prevalence of SARS-CoV-2 antibodies of 11.0% (n=33). Of these, 13 (39.4%) presented both IgM and IgG, 18 (54.5%) only IgG, and 2 (6.1%) only IgM. For the ELISA test, which measures only IgG and not IgM, and considering the result as a categorical variable according to the manufacturer’s threshold described in the methods, the serological prevalence was 13.0% (n=39), slightly higher than with the LFA test (Table 1). Considering the greater sensitivity that ELISA tests have over LFA tests^22^ and the two cases from the LFA tests that were positive for IgM only (which is not detected by this ELISA test), we obtain for this population a total serological prevalence of SARS-CoV-2 infection of 13.7% (n=41 positives to IgG or IgM). Based on the contacts reported by participants with positive results for the ELISA or LFA tests (n=41), we estimate that 15 cases (36.6%) were likely to be infected outside the hospital, 14 cases (34.1%) inside, and in 12 cases (29.3%) there is insufficient information to estimate. Of these 41 positive subjects, 9 individuals (22.0%) reported working additional shifts in other hospitals, 4 of them (9.8%) in a COVID hospital, and 5 (12.2%) in other non-COVID hospitals. Within these same 41 positive subjects, close to half (n=19, 46.3%) reported the day of the blood sample not having experienced related symptoms, nor suspected having had COVID-19 (Table 1).

### Test comparisons

From our sample of 300 participants, 87 (29.0%) reported they had previously had RT-PCR tests for SARS-CoV-2, 10 of them having positive results (11.5% positivity rate). The median number of days between the date of the RT-PCR tests that were positive and when the blood sample was taken in these 10 individuals was 93 days (range, 48-117 days). The concordance between the 10 RT-PCR positive subjects and the ELISA test was 8 (80.0%), slightly higher than with the LFA test which was 7 (70.0%). Compared to the ELISA test, the LFA test had sensitivity of 79.5%, specificity of 100.0%, positive predictive value of 100.0% and negative predictive value of 97.0%.

### Factors associated with SARS-CoV-2 infection

Remarkably, the security and janitorial work groups had substantially higher rates of positive results (62.5% and 45.4%, respectively) than the other work groups (Table 1). Additionally, participants with educational level below university degree also had higher rates of positive results than those with a higher educational level (23.5% vs. 9.2%). The variables that presented *β* coefficients with the strongest associative strength to the level of anti-SARS-CoV-2 IgG antibodies are: working in the security group, educational level, the number of persons with which one lives, and the following symptoms: muscle and joint pain, dyspnea, fever, conjunctivitis, olfactory alterations and dysgeusia (Table 2).

**Table 2.**
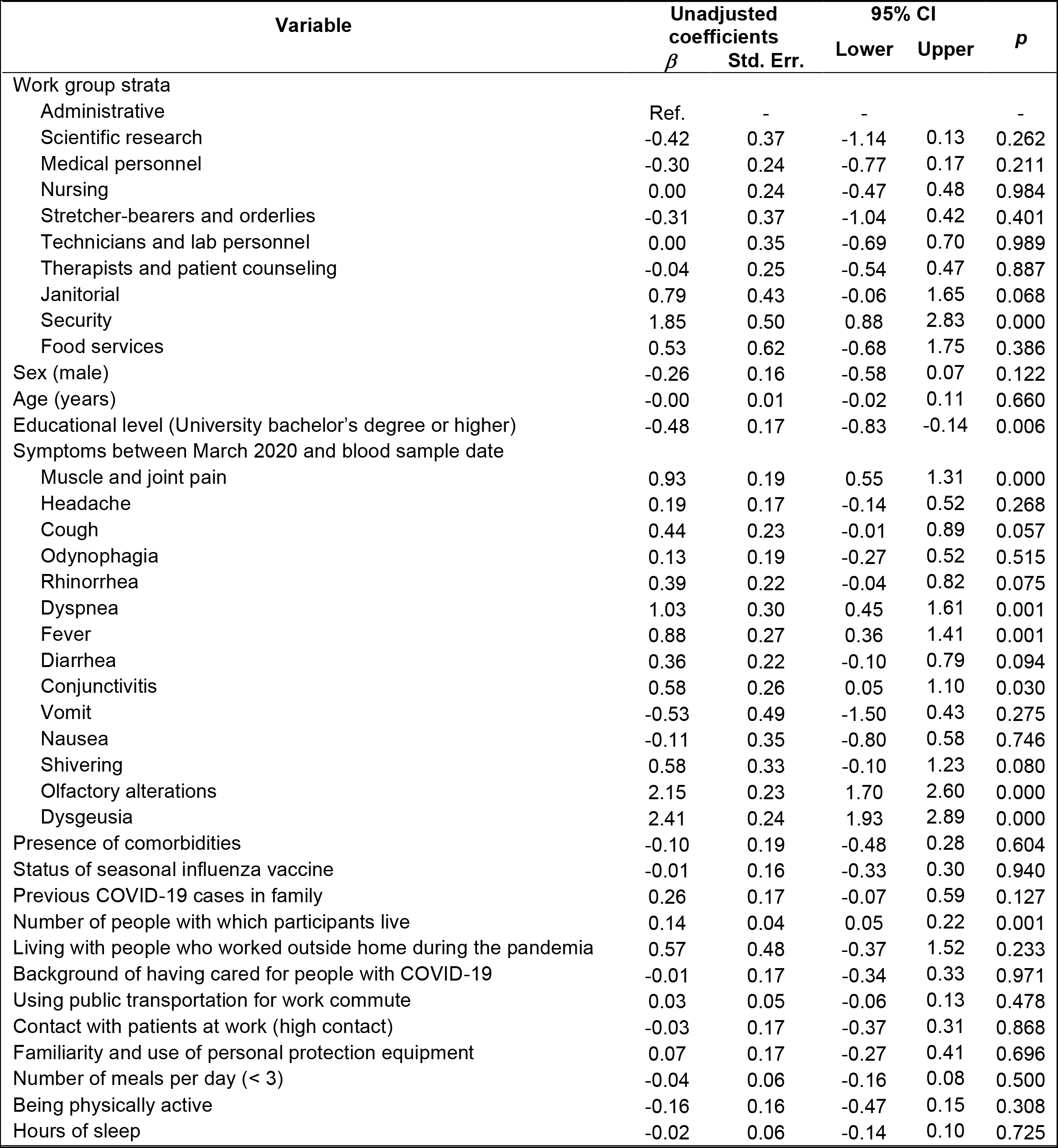
Linear regression analysis of associations between variables and result ratios from ELISA tests.

| Variable | Unadjusted coefficients |  | 95% CI |  | p |
| --- | --- | --- | --- | --- | --- |
| | $\beta$ | Std. Err. | Lower | Upper | |
| Work group strata |  |  |  |  |  |
| Administrative | Ref. | - | - | - | - |
| Scientific research | -0.42 | 0.37 | -1.14 | 0.13 | 0.262 |
| Medical personnel | -0.30 | 0.24 | -0.77 | 0.17 | 0.211 |
| Nursing | 0.00 | 0.24 | -0.47 | 0.48 | 0.984 |
| Stretcher-bearers and orderlies | -0.31 | 0.37 | -1.04 | 0.42 | 0.401 |
| Technicians and lab personnel | 0.00 | 0.35 | -0.69 | 0.70 | 0.989 |
| Therapists and patient counseling | -0.04 | 0.25 | -0.54 | 0.47 | 0.887 |
| Janitorial | 0.79 | 0.43 | -0.06 | 1.65 | 0.068 |
| Security | 1.85 | 0.50 | 0.88 | 2.83 | 0.000 |
| Food services | 0.53 | 0.62 | -0.68 | 1.75 | 0.386 |
| Sex (male) | -0.26 | 0.16 | -0.58 | 0.07 | 0.122 |
| Age (years) | -0.00 | 0.01 | -0.02 | 0.11 | 0.660 |
| Educational level (University bachelor's degree or higher) | -0.48 | 0.17 | -0.83 | -0.14 | 0.006 |
| Symptoms between March 2020 and blood sample date |  |  |  |  |  |
| Muscle and joint pain | 0.93 | 0.19 | 0.55 | 1.31 | 0.000 |
| Headache | 0.19 | 0.17 | -0.14 | 0.52 | 0.268 |
| Cough | 0.44 | 0.23 | -0.01 | 0.89 | 0.057 |
| Odynophagia | 0.13 | 0.19 | -0.27 | 0.52 | 0.515 |
| Rhinorrhea | 0.39 | 0.22 | -0.04 | 0.82 | 0.075 |
| Dyspnea | 1.03 | 0.30 | 0.45 | 1.61 | 0.001 |
| Fever | 0.88 | 0.27 | 0.36 | 1.41 | 0.001 |
| Diarrhea | 0.36 | 0.22 | -0.10 | 0.79 | 0.094 |
| Conjunctivitis | 0.58 | 0.26 | 0.05 | 1.10 | 0.030 |
| Vomit | -0.53 | 0.49 | -1.50 | 0.43 | 0.275 |
| Nausea | -0.11 | 0.35 | -0.80 | 0.58 | 0.746 |
| Shivering | 0.58 | 0.33 | -0.10 | 1.23 | 0.080 |
| Olfactory alterations | 2.15 | 0.23 | 1.70 | 2.60 | 0.000 |
| Dysgeusia | 2.41 | 0.24 | 1.93 | 2.89 | 0.000 |
| Presence of comorbidities | -0.10 | 0.19 | -0.48 | 0.28 | 0.604 |
| Status of seasonal influenza vaccine | -0.01 | 0.16 | -0.33 | 0.30 | 0.940 |
| Previous COVID-19 cases in family | 0.26 | 0.17 | -0.07 | 0.59 | 0.127 |
| Number of people with which participants live | 0.14 | 0.04 | 0.05 | 0.22 | 0.001 |
| Living with people who worked outside home during the pandemia | 0.57 | 0.48 | -0.37 | 1.52 | 0.233 |
| Background of having cared for people with COVID-19 | -0.01 | 0.17 | -0.34 | 0.33 | 0.971 |
| Using public transportation for work commute | 0.03 | 0.05 | -0.06 | 0.13 | 0.478 |
| Contact with patients at work (high contact) | -0.03 | 0.17 | -0.37 | 0.31 | 0.868 |
| Familiarity and use of personal protection equipment | 0.07 | 0.17 | -0.27 | 0.41 | 0.696 |
| Number of meals per day (< 3) | -0.04 | 0.06 | -0.16 | 0.08 | 0.500 |
| Being physically active | -0.16 | 0.16 | -0.47 | 0.15 | 0.308 |
| Hours of sleep | -0.02 | 0.06 | -0.14 | 0.10 | 0.725 |

Table 3 shows two adjusted linear regression models using the ratio results of the ELISA test. Both show that male participants had lower levels of IgG antibodies than female participants. Reporting muscle and joint pain, and olfactory alterations were significantly associated with increments in antibody levels. The level of antibodies also increased with the number of people with which participants live. Model 1 shows additionally that the janitorial (*β* = 1.02, 95% confidence interval [CI], 0.28-1.76) and security (*β* = 1.10, 95% CI, 0.23-1.97) work groups were strongly associated with the ELISA results. Exchanging the work group variable with the educational level in Model 2, all remaining variables present very similar associations than those in Model 1, with the additional result that participants with educational level of a university bachelor’s degree or higher had a reduction in the results of the ELISA test (*β* = −0.36, 95% CI, −0.67, −0.06) compared to those participants with lower educational levels.

**Table 3.**
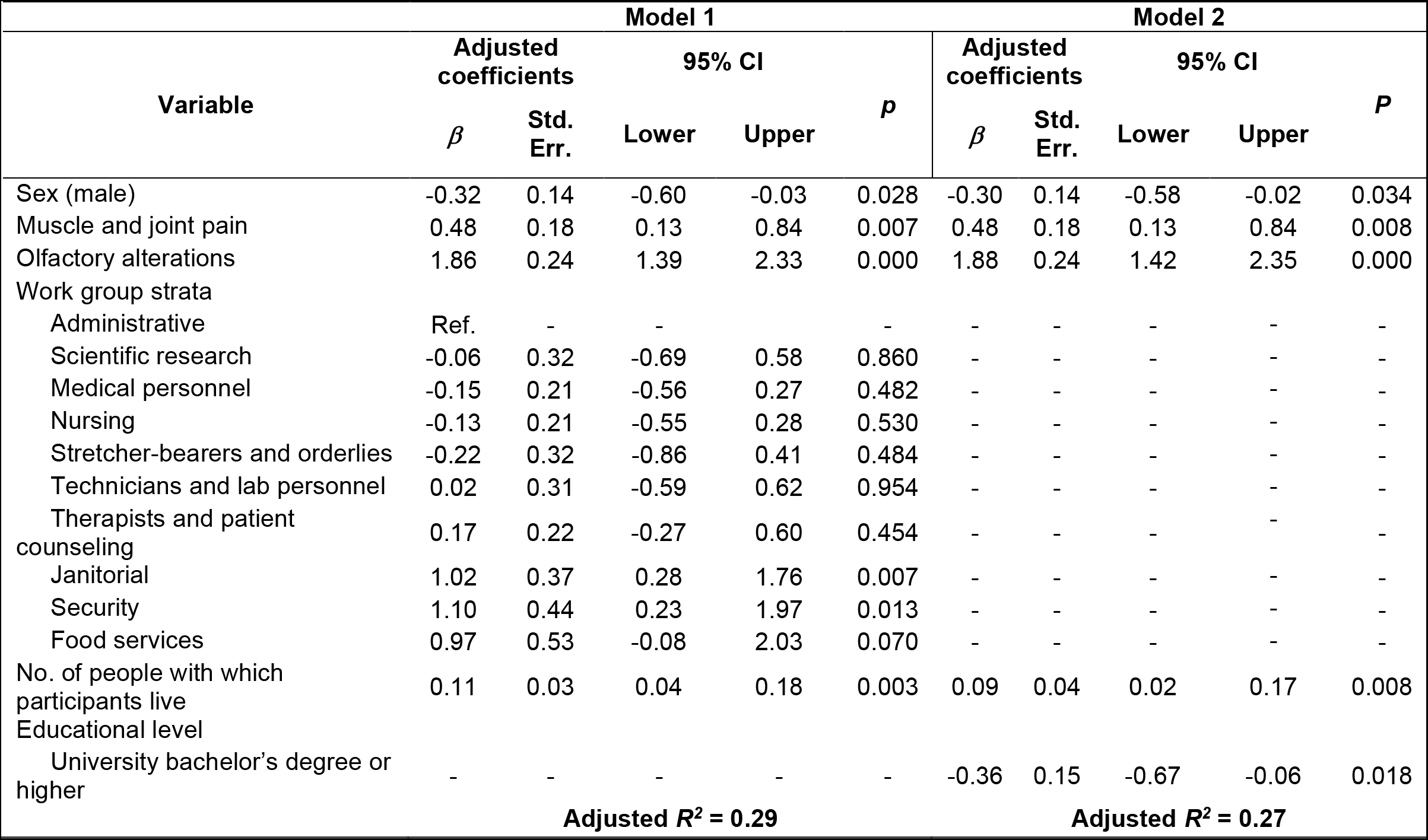
Linear regression models between associated variables and result ratios from ELISA tests.

## DISCUSSION

In this work we provide results from a seroprevalence survey in a large non-COVID teaching hospital in Mexico City. The ELISA results indicate that 13.0% of the sample are positive to IgG antibodies to SARS-CoV-2. Considering also the results from the LFA tests which identified additional subjects positive for IgM but negative for IgG antibodies, we obtain an overall prevalence in our sample of SARS-CoV-2 antibodies of 13.7%. We consider this last estimate to more closely reflect the situation since it combines the better sensitivity of the ELISA test, with the additional IgM information provided by the LFA test. Our study also identified several associated sociodemographic factors that increase the risk of having previous infection with SARS-CoV-2: working in the janitorial and security groups, having an education level below university bachelor’s degree, and living with a larger number of people.

A study with HCW in London found that 21.9% of seropositive subjects did not report COVID-19 symptoms^14^. In this work, almost half (46.3%) of seropositive subjects had experienced no symptoms and did not suspect they had previously had SARS-CoV-2 infection. These numbers underscore the importance of seroprevalence surveys including asymptomatic subjects as fundamental in order to obtain accurate information regarding the extent of infections in a population.^17,23^

Recent studies have found that seropositivity of antibodies to SARS-CoV-2 decreases substantially over a 60-day period of time^21^. Additionally, it is unclear whether all patients with COVID-19 elevate antibodies, and in which time frame^18,19,20,24^. This implies that in our current study we may not be capturing all the subjects that might have been previously infected with SARS-CoV-2, and therefore our result of 13.7% prevalence of SARS-CoV-2 should be considered as a lower bound in terms of the prevalence of previous infection to this virus. Another point that must be considered is that given that the questionnaire and blood samples were taken the same day, we do not have precise knowledge of the temporal sequence of events, and therefore a cohort study would provide more clarity in this regard.

Our linear regression model based on the ELISA test indicates that male subjects tend to have lower antibody scores than female subjects. This is in line with previous studies finding that in some groups of COVID-19 patients the generation of IgG antibodies is stronger in women than men^25^ and that in general women mount stronger immune responses to infections and vaccinations.^26^

Compared with studies measuring seropositivity for HCW in other countries, the antibody prevalence observed in our study for the whole hospital (13.7%) is within the range of that found for other countries: 19.1% for a large hospital in Sweden^13^, 10.2% for nationwide HCW in Spain^9^ and 6.2% for a tertiary care center in Belgium^16^. Since our hospital was designated non-COVID, it is to be expected to have fewer infections than in a hospital that was focused on COVID-19 patients (however, 9.8% of the positive cases in our study reported working other shifts in a COVID hospital).

Additionally, there is still patient contact in our hospital, and some patients that later were known to have positive RT-PCR tests were treated by the subjects in our sample. Thus, in a sense it is more representative of the general population, as well as other work environments where SARS-CoV-2 positive individuals occur randomly, rather than being specifically grouped as in a COVID-19 hospital.

In addition to physiological factors that are known to be associated with COVID-19, such as olfactory alterations, fever, muscle and joint aches, we find that the prevalence of antibodies is associated with three sociodemographic factors: number of people with which one lives, having an educational level below a university bachelor’s degree, and type of work, with subjects working as janitors and security guards having substantially higher risk than other personnel groups such as physicians, nurses and administrative staff (Table 3). Both linear regression models we present here each reflect different explicative variables: Model 1 with an emphasis of work groups within the hospital and Model 2 of a socioeconomic factor that goes beyond their work in the hospital. Thus, each model reflects different aspects of sociodemographic factors that imply that in this population, less favored socioeconomic groups, i.e. those with lower educational level and living with more people, are at significantly higher risk of being infected with SARS-CoV-2. Interestingly, another seroprevalence study among HCW from the New York City area^15^ also found that service/maintenance personnel (including, housekeepers and groundkeepers, among others) had a substantially larger level of SARS-CoV-2 exposure (20.9%) than other professions such as physicians (8.7%) and allied health professionals such as physical and occupational therapists (11.6%). This suggests that the sociodemographic factors identified in our seroprevalence survey may also be associated with higher SARS-CoV-2 exposure in other contexts, including other countries.

Our sample can be representative of other non-COVID hospitals in other regions in Mexico with similar degrees of contagion to Mexico City. Additionally, given the filters limiting access to the hospital to COVID-19 patients, in the absence of other data it might also provide relevant information to other non-healthcare institutions or organizations with similar sociodemographic and educational breakups as our institution. Seroprevalence studies in various populations can be a tool to provide useful information for planning public health measures at institutional, regional and national levels. By September 9^th^, which is the date our last blood sample was collected, the number of confirmed cases in Mexico City was 107,613 which represented 1.2% of the city’s population. Given the sentinel model of epidemiological surveillance that has been followed by the Mexican authorities, the more than ten-fold difference between this number and the 13.7% obtained in our study is not surprising and cannot be accounted for as a difference caused by risks specific to HCWs. Although not representative at a regional or national level, in the current absence of other data, this seroprevalence survey provides useful information that can give a sense of the extent of the pandemic in a specific segment of the Mexican population and contribute valuable information to guiding public health policies.

## METHODS

### Study population

We performed a cross-sectional study, having as study population the personnel that works in the hospital (n=3148). We assumed a positivity rate of 10% in the general population, and a significance level of 5%, with which we calculated a sample size of 139. In order to increase the sample power, and given the lack of information about COVID-19 prevalence in the Mexican population, we selected a final sample size of 300 subjects. The subjects in the sample were stratified in 10 groups according to their work activities: administrative; scientific research; medical personnel; nursing; stretcher-bearers and orderlies; technicians and laboratory personnel; therapists and patient counseling; janitorial; security; and food services. We selected participants randomly from each of the 10 work group strata, according to the size of each. Inclusion criteria were: being personnel working in the hospital, from any shift, and granting written informed consent. Participants were contacted by telephone, those that did not consent to participate in the study were replaced by other participants selected by the same randomized process. The study was approved by the hospital’s Ethics and Research Committees (approval number INR-26/20, approval date July 6^th^, 2020) and was carried out in accordance with relevant guidelines and regulations, including the Declaration of Helsinki. Written informed consent was obtained from all participants.

### Data collection

We scheduled selected participants between August 10^th^ and September 9^th^, 2020, to obtain a peripheral blood sample and complete a questionnaire. We conducted procedures in a specially designated area, following World Health Organization safety guidelines with both participants and study personnel: use of facemasks, physical distancing, ventilated area, hand hygiene, etc.

### Definition of variables

In the questionnaire, we explored sociodemographic variables, COVID-19 previous or present signs and symptoms, diagnostic RT-PCR tests, comorbidities, status of seasonal influenza vaccine, living arrangements (including number of people sharing a home, family relationship, number of bedrooms), means of transportation, degree of exposure to patients, use of personal protective equipment, number of meals per day, physical activity, and hours of sleep. With the blood samples, we conducted two serological tests, an LFA and an ELISA. The LFA tests were the COVID-19 IgG/IGm Rapid Test Cassette® (Hangzhou Biotest Biotech Co. Ltd., China), which use an immunochromatographic assay to provide qualitative detection of IgM and IgG antibodies against the SARS-CoV-2 virus. We considered a participant to be positive with the LFA test if they had a positive result for either IgG, IgM or both. The ELISA tests we used were the Euroimmun® Anti-SARS-CoV-2 NCP which detect IgG antibodies against the SARS-CoV-2 nucleocapsid protein (Euroimmun Medizinische Labordiagnostika, AG, Germany). According to the manufacturer’s guidelines, a positive result is one with a resulting ratio ≥ 1.1, negative ≤ 0.8 and the interval between these values is considered uncertain. The ratio which results from this test was considered a non-categorical continuous variable for the linear regression models.

### Statistical analysis

We conducted all statistical analyses with Stata v13.0 (StataCorp, USA). We report proportions of sample study groups and qualitative variables as percentages. We used linear regression analysis to obtain unadjusted *β* coefficients, standard errors (Std. Err.), 95% confidence intervals (CI), and *p* values, in order to evaluate associations between the variables and the resulting ratios of the ELISA tests. Those associations with *p* values ≤ 0.25 were selected to adjust linear regression models. Each variable was then incorporated into the model and those that had *p* ≤ 0.05 were retained. We tested with an F statistic the fitting of the reduced models with an adjusted R^2^. Finally, and using the thresholds established by the ELISA test manufacturer, we calculated the sensitivity, specificity, positive and negative predictive values of the LFA.

### Data availability

**T**he data supporting the current study has not been deposited in a public repository because of confidentiality concerns, but sharable data can be made available upon reasonable request.

## Data Availability

Data referred to in the mansucript can be made available by contacting the corresponding author.

## Acknowledgments

The authors would like to thank José Ovidio Marín Salcedo and Manuel Martínez Torres for their generous support providing the serological tests. We are also grateful to Luis Esaú López Jácome, Guillermo Cerón González and Rocío Vázquez Juárez for their assistance in collecting the blood samples.

## Author contributions

J.P.O. and C.I. conceived the work. J.P.O., C.I., E.C.A., T.Ch., S.L.B., C.A.C.C., and R.F.C. designed the study. E.C.A., E.C.R., S.L.B., C.A.C.C., and T.Ch. executed the study and collected the data. E.C.A., J.P.O., C.A.C.C., and E.C.R. analyzed the data. J.P.O., E.C.A., E.C.R., and R.F.C. drafted the work. All authors provided input into the final manuscript.

